# N-Acetylcysteine Reduces Tryptophan-induced Abnormalities in People with Schizophrenia

**DOI:** 10.64898/2026.06.25.26356572

**Authors:** Stephanie M. Hare, Deanna L. Kelly, Yezhi Pan, Shuo Chen, Frank Blatt, David A. Gorelick, James M. Gold, Korrapati V. Sathyasaikumar, Bhim M. Adhikari, Peter Kochunov, S. Andrea Wijtenburg, Laura M. Rowland, Robert Schwarcz, Robert W. Buchanan

## Abstract

The current study assessed whether N-acetylcysteine (NAC), which inhibits the kynurenic acid (KYNA)-synthesizing enzyme kynurenine aminotransferase (KAT) II, affects tryptophan (TRYP)-induced peripheral formation of the kynurenine pathway metabolites kynurenine and KYNA and improves selected functional outcome measures in people with schizophrenia. Fifty-eight participants with DSM-5 schizophrenia or schizoaffective disorder entered a double-blind, placebo-controlled, randomized cross-over challenge study, in which they were pretreated with either NAC (up to a maximum of 15 g) or placebo, then received TRYP, 6 g. Prior to and after receiving the study medications, participants underwent laboratory (serum kynurenine and KYNA), symptom (BPRS, SANS, and CDS), cognitive (6 MCCB tests) and brain MRI (ASL, DTI, ^1^H-MRS) assessments. In contrast to placebo pre-treatment, NAC significantly reduced the TRYP-induced increase in peripheral serum levels of kynurenine (t=-2.02; p<0.05) and KYNA (t=-3.21; p=0.002). NAC pre-treatment was associated with significantly smaller increases in total white matter (WM) cerebral blood flow (CBF) (t=-2.15; p=0.04) and a trend for smaller increases in total gray matter (GM) CBF (t=-1.81; p=0.08). NAC pre-treatment significantly reduced the TRYP-induced decrease in MCCB composite score (t=2.07; p=0.04). There was no differential treatment effect on DTI or ^1^H-MRS or symptom measures. The observation that NAC attenuated the *de novo* formation of KYNA, reduced WM CBF elevations, tended to decrease GM CBF, and blocked the worsening of cognitive performance in participants following TRYP administration, supports the concept that KAT II inhibition is a promising novel strategy for the treatment of cognitive impairments in people with schizophrenia.

## Introduction

People with schizophrenia (PSz) are characterized by marked cognitive impairments, which are major determinants of poor social and occupational outcomes [1,2]. Antipsychotic medications have limited benefits for these impairments [3,4], and studies of add-on pharmacological agents for their treatment have been largely disappointing [5–10]. In the absence of effective treatments, cognitive impairments remain a critical unmet therapeutic need.

There is converging evidence to suggest that disturbances in the kynurenine pathway (KP) [11], the major route of tryptophan (TRYP) metabolism, are related to the pathophysiology of schizophrenia. Clinical studies of PSz demonstrate increased levels of kynurenine and kynurenic acid (KYNA) in cerebrospinal fluid [12–14], while post-mortem studies have found increased levels of kynurenine and KYNA in the prefrontal cortex [14–17]. There are also documented changes in two KP enzymes: tryptophan 2,3-dioxygenase (TDO) [16,18], and kynurenine 3-monooxygenase (KMO) [17], with the changes in enzyme activity leading to increased KYNA production.

Genetic studies provide further support for dysfunction of the KP in PSz. The KMO single nucleotide polymorphism (SNP): rs2275163 C risk allele is associated with decreased KMO gene expression in the frontal eye field and impaired predictive eye pursuit and visual working memory in PSz [19]. In a second study, Wonodi et al found that this risk allele was associated with impaired neuropsychological test performance in PSz and there was a trend for the KMO SNP rs1053230 C risk allele to be associated with impaired cognitive function [20]. Holtze et al found an association between this latter SNP and CSF KYNA levels [21].

In the mammalian central nervous system (CNS), KYNA is a known antagonist of the α-7 nicotinic and N-methyl-D-aspartate (NMDA) glutamate receptors [22]. Altered KYNA antagonism of these receptors is hypothesized to be a critical mechanism in the development of the cognitive impairments observed in schizophrenia [11]. The conversion of kynurenine to KYNA in the CNS is controlled primarily by the enzyme kynurenine aminotransferase (KAT II). In animal models, inhibition of KAT II improves cognitive function by decreasing KYNA levels in the CNS [11]. Unfortunately, there are currently no KAT II inhibitors available for human commercial use.

N-acetylcysteine (NAC) is an acetylated derivative of L-cysteine, with multiple pharmacological properties including glutamatergic modulation and putative neurotrophic and anti-inflammatory effects [23]. In addition, NAC is a precursor of the antioxidant glutathione [23]. Moreover, we have recently demonstrated that NAC inhibits KAT II activity. Specifically, we found that NAC dose-dependently: 1) reduces the neosynthesis of KYNA from kynurenine in rat and human brain tissue homogenates and in rat brain tissue slices; and 2) inhibits the activity of recombinant human KAT II (IC_50_: ∼500 µM) [24]. In addition, NAC pre-treatment dose-dependently attenuates the *de novo* production of KYNA from kynurenine *in vivo*, significantly reducing extracellular KYNA in the rat prefrontal cortex [24]. These results provide strong support for the idea that KAT II inhibition by NAC can reduce cerebral KYNA levels in humans.

In a previous study, we administered TRYP to PSz and healthy controls to examine its effects on plasma concentrations of kynurenine and KYNA and on neuroimaging and cognitive outcome measures [25]. As predicted, the TRYP challenge produced significant, time-dependent elevations in plasma kynurenine and KYNA in both cohorts. Furthermore, cerebral blood flow (CBF) was affected differentially, such that TRYP versus placebo administration was associated with significantly higher brain gray matter CBF in the healthy controls [25]. Although TRYP did not significantly impair cognitive test performance, there was a trend for TRYP to worsen visuospatial memory task performance in the healthy controls [25]. These results demonstrated that TRYP substantially increases plasma levels of kynurenine and KYNA in both groups, but exerts differential group effects on CBF.

The current study was designed to examine whether high-dose NAC reduces the TRYP-induced elevation of peripheral KYNA and the effects associated with increased KYNA on selected measures of brain behavior and function, including CBF, ^1^H-MRS glutamate and glutathione, and neuropsychological test performance. The demonstration that KAT II inhibition leads to beneficial effects on cognitive function would represent a major development in the therapeutics of schizophrenia.

## Materials and Methods

### Inclusion/Exclusion Criteria

Participants were either male or female, of any race and ethnicity, between 18-55 years old, and met Diagnostic and Statistical Manual of Mental Disorders 5^th^ Edition (DSM 5) [26] criteria for schizophrenia or schizoaffective or schizophreniform disorder. A best estimate diagnostic approach was utilized in which information from the Structured Clinical Interview for DSM 5 [27] was supplemented by information from family informants and medical records to generate a diagnosis. Sex, race and ethnicity were ascertained through participant interview. Participants were treated with either first or second generation antipsychotic medication and on a stable dose of the medication(s) for 30 days prior to study entry.

Exclusion criteria included current DSM 5 substance use disorder other than nicotine or mild cannabis use disorders; medical disorders that have known effects on TRYP metabolism/absorption; current treatment with monoamine oxidase inhibitors, triptans, and dextromethorphan; history of a brain disorder whose pathology could alter the presentation or treatment of schizophrenia or significantly increase the risk associated with the proposed treatment protocol; daily caffeine intake greater than 1000 mg; contraindication for MRI; and currently pregnant or breastfeeding.

The University of Maryland School of Medicine IRB approved the study protocol and informed consent procedures. Written informed consent was obtained from all participants after the study procedures had been fully explained and prior to study participation. The ability to provide valid informed consent was documented using an evaluation to sign consent for all study participants. The study was registered with clinicaltrials.gov (NCT04013555).

### Clinical Assessments

The Brief Psychiatric Rating Scale (BPRS) [28] was used to assess global psychopathology and positive symptoms; the Scale for the Assessment of Negative Symptoms (SANS) [29] was used to assess negative symptoms; Calgary Depression Scale (CDS) [30] was used to assess depressive symptoms; and the Clinical Global Impression Scale (CGI) [31] severity of illness item was used to assess global changes.

### Cognitive Assessments

The assessment consisted of six tests drawn from the MATRICS™ Consensus Cognitive Battery (MCCB) [32]: the Hopkins Verbal Learning Test Revised (HVLT-R), the Letter-Number Sequencing (LNS) test, the Continuous Performance Test - Identical Pairs (CPT-IP), the Brief Visuospatial Memory Test Revised (BVMT-R), the Working Memory III Spatial Span (WMSIII-SS) Test, and the BACS Symbol Coding (BACSS) Test. The six tests were combined to form a composite score.

### Safety Assessments

A standard blood chemistry panel, complete blood count, urinalysis, urine pregnancy test (for female participants of child-bearing potential) and EKG were obtained during the Evaluation Phase. On each challenge day, the participant underwent baseline and post-treatment vital sign and side effect evaluations, documented with the Side Effect Checklist (SEC). The SEC is comprised of 32 common side effects, which are rated on a 1 (none)-4 (severe) Likert scale.

### Kynurenine and KYNA Measurements

Peripheral venous blood samples were collected in EDTA-containing tubes and centrifuged within 30 min after the blood draw. After centrifugation, the supernatant plasma was removed and stored at -80°C until analysis. Detailed description of the assay procedures is presented in the Supplementary Methods.

### SNP Genotyping

Genomic DNA was extracted from whole blood collected in PAXgene Blood DNA Tubes using the PAXgene Blood DNA Kit (QIAGEN/PreAnalytiX, Cat. No. 761133) according to the manufacturer’s instructions. A total of 10 ng of purified genomic DNA was used for genotyping. We focused primarily on the *KMO* variants rs2275163 (C>T) and rs1053230 (C>T). Both SNPs have minor allele frequencies ≥ 0.15 in European American and African American/mixed populations. Detailed description of the genotyping procedures is presented in the Supplementary Methods.

### MRI Assessments

The following MRI scans were obtained using a Siemens PRISMA^fit^ MR system with a 64-channel phased array head/neck coil: structural-T1; arterial spin labeling (ASL); magnetic resonance spectroscopy (^1^H-MRS); and diffusion tensor imaging (DTI). Detailed description of the neuroimaging procedures is presented in the Supplementary Methods.

### Study Medications

The NAC powder was obtained from Medisca Inc. (Plattsburgh, NY USA). NAC was dispensed in a 12 oz amber bottle, which contained the following ingredients: NAC (140 mg/kg body weight up to a maximum of 15 g total); maltodextrin, 60 g, obtained from the Bulk Supplement Manufacturer (Henderson, NV USA); sucrose, 30 g, from Domino Foods Inc. (West Palm Beach, FL USA); and sodium bicarbonate USP, 6 g, from Medisca Inc. The placebo contained maltodextrin, 60 g; sucrose, 15 g; and sodium bicarbonate, 6 g. The NAC and placebo dry powder was diluted with 300 ml of distilled water at the time of dispensation.

The TRYP powder was obtained from Ajinomoto, North America Inc. (Eddyville, IA, USA) and was approved for use in the trial by the FDA (IND# 114931). The TRYP slurry was prepared by mixing 6 g of TRYP with 7.8 g of Domino 10X Confectioner’s Sugar (Yonkers, NY, USA) and 6 g of Nestlé’s Cocoa mix (Glendale, CA, USA). The dried mixture was placed into an 8 oz amber bottle, and the mixture was dissolved in 150 ml of distilled water.

The study participants were instructed to drink each solution within a one-minute period. The study medications were dispensed by a research pharmacist who did not participate in assessment procedures.

### Study Design

The study used a randomized, double-blind, crossover design, in which participants were pre-treated with either oral high-dose NAC or placebo, then received an oral dose of TRYP, 6 g. The study was comprised of two phases: an Evaluation Phase and a Challenge Phase. In the Evaluation Phase, all participants signed informed consent and completed screening and baseline medical and safety assessments. The Challenge Phase was comprised of two challenge days: each challenge day included a morning fasting blood draw; and baseline MRI, neuropsychological and clinical assessments. After completion of the MRI scan, the participant was administered either NAC or placebo, then 30 minutes later TRYP. Protein-free snacks were provided shortly after TRYP ingestion. The second MRI assessment was conducted 90 minutes after TRYP and the second neuropsychological and clinical assessments were conducted 140-180 minutes after TRYP administration. There were three post-baseline blood draws: 60, 140 and 240 minutes after TRYP administration. The two challenge days were at least 2 weeks apart. Randomization used a permuted block design of 4, stratified by sex.

### Statistical Analysis

To evaluate the NAC effect, we used intra-subject contrasts of the outcomes. Specifically, difference-in-difference scores were calculated for each subject, defined as the difference in post- vs. pre-challenge outcomes on the NAC challenge day, minus the post- vs. pre-challenge outcomes on the placebo challenge day. This approach was chosen because it more effectively adjusts for both measured and unmeasured time-invariant confounding factors. We also assessed potential carryover effects inherent to the crossover design and adjusted for them as necessary. A statistically significant deviation of the intra-subject contrast from zero was interpreted as evidence of an NAC effect.

For the analysis of peripheral KYNA and kynurenine levels (primary aim 1), both metabolites were measured at multiple time points on each challenge day (see **Study Design**); we employed linear mixed-effects models including time as a categorical covariate. This approach allowed us to assess whether the NAC effect was significant at specific time points.

For the analyses of imaging measures including ASL (primary aim 2), DTI and MRS (primary aim 3), and exploratory analyses of clinical and cognitive outcomes, we fit separate linear regression models adjusting for the randomization sequence (i.e., whether a participant received NAC first or placebo first).

As a final (additional) exploratory analysis, we examined the effect of the two KMO SNP C risk alleles on outcome measures for which there was a significant effect of NAC treatment. We used the same linear regression model as described above for these analyses.

All hypothesis tests were two-sided, with a significance threshold of p < 0.05.

## Results

Eighty-eight people were consented, of whom 70 were eligible and randomized to study treatments (see Supplementary Figure 1: CONSORT Flow Chart). Fifty-one participants completed both challenge days. Of the 19 participants who were withdrawn from the study prior to completion, 7 completed at least one challenge day and were included in analyses. See Supplementary Table 1 for the demographic data of these 58 participants.

**Table 1:** Kynurenine and Kynurenic Acid (KYNA) Plasma Levels.

| Kynurenine (pmoles/μl) |  |  |  |  |
| --- | --- | --- | --- | --- |
| Timepoint | N-acetylcysteine |  | Placebo |  |
|  | mean | ± SD | mean | ± SD |
| Baseline | 2.39**** | 1.89 | 1.97*** | 1.22 |
| 60 minutes | 22.0* | 18.4 | 23.3**** | 37.3 |
| 240minutes | 65.8** | 51.6 | 94.0** | 159.5 |
| KYNA (fmoles/μl) |  |  |  |  |
| Timepoint | N-acetylcysteine |  | Placebo |  |
|  | mean | ± SD | mean | ± SD |
| Baseline | 48.1**** | 50.9 | 42.9*** | 35.2 |
| 60 minutes | 1687*** | 3463 | 1322**** | 3382 |
| 240 minutes | 5758** | 4850 | 7471** | 6930 |

\*: n=51; \*\*: n=52; \*\*\*: n=53; \*\*\*\*: n=54

### Kynurenine and KYNA

In the analyses examining the effect of NAC compared to placebo on kynurenine and KYNA plasma levels, we only included the baseline, 60 minute and 240 minute samples, because of extensive missing data for the 140 minute timepoint (see Table 2). There was a significant effect of NAC on both kynurenine (t=-2.02; p=0.048) and KYNA (t=-3.21; p=0.002) levels at 240 minutes (see Figure 1 and Table 1). The difference at 60 minutes was not significant for kynurenine (t=-0.06; p=0.96) nor KYNA (t=-0.66; p=0.51).

**Figure 1:**
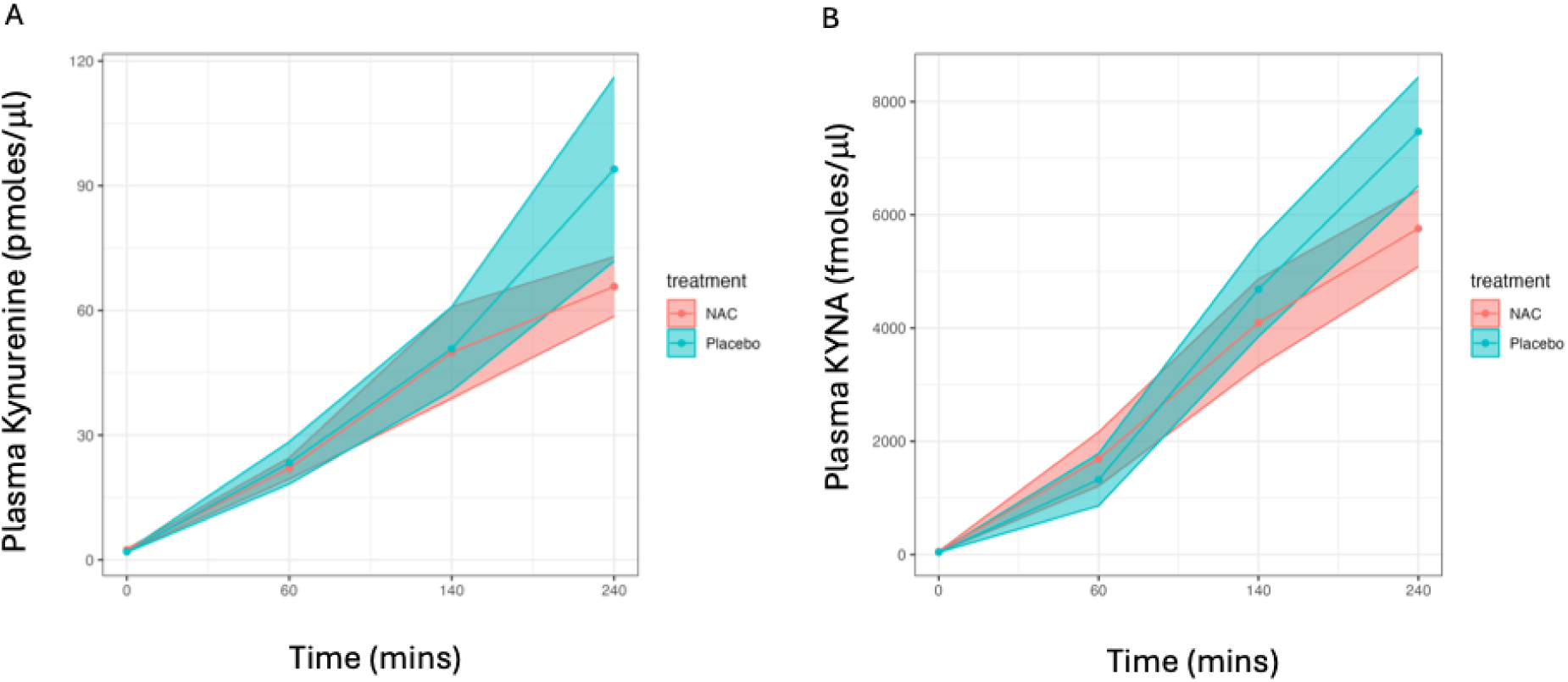
Effects of NAC on Tryptophan-Induced Elevations of Kynurenine and KYNA. Footnote: ^a^The shaded areas represent 95% confidence intervals for kynurenine (Panel A) and KYNA (Panel B) over the course of sample collection. There was a significant effect of NAC on both kynurenine (t=-2.02; p=0.048) and KYNA (t=-3.21; p=0.002) levels at 240 minutes. ALT text: The shaded areas represent 95% confidence intervals for kynurenine and KYNA over the course of sample collection. There was a significant effect of NAC on kynurenine and KYNA levels at 240 minutes.

### Neuroimaging Measures

<u>ASL</u>. There were significant carry-over effects for multiple brain regions, therefore all analyses included order of NAC administration in the model. There was a significant effect of NAC compared to placebo pre-treatment on the pre-/post-treatment change in average whole brain white matter CBF (avg-WM CBF; t=-2.151; p= 0.037), with a trend for a significant effect of NAC pre-treatment on the pre-/post-treatment change in average whole brain gray matter CBF (avg-GM CBF; t=-2.15; p=0.078). For both avg-WM and avg-GM CBF, NAC pretreatment reduced the CBF elevations that were observed following TRYP administration (see Table 2 and Figure 2).

In exploratory analyses, we examined the change in gray matter CBF in cortical (n=210) and subcortical (n=36) regions defined by the Brainnetome Atlas [33]. In 5 cortical regions, there was a difference in CBF with a Cohen’s d effect size of 0.50 or greater. In an additional 55 cortical regions, there was a difference in CBF with a Cohen’s d effect size between 0.30 and 0.50. In all cases, NAC reduced the increase in CBF following TRYP administration compared to the placebo arm. In 27 of the 36 subcortical regions, NAC treatment reduced the increase in CBF following TRYP administration, but none of the treatment differences had a Cohen’s d effect size greater than 0.32. The cortical regions with a medium effect size or greater clustered in regions that have been previously implicated in the neuroanatomy of schizophrenia: superior temporal gyrus, posterior temporal sulcus, inferior parietal lobule, insular gyrus and cingulate gyrus [34] (see Supplementary Figure 2).

**Figure 2:**
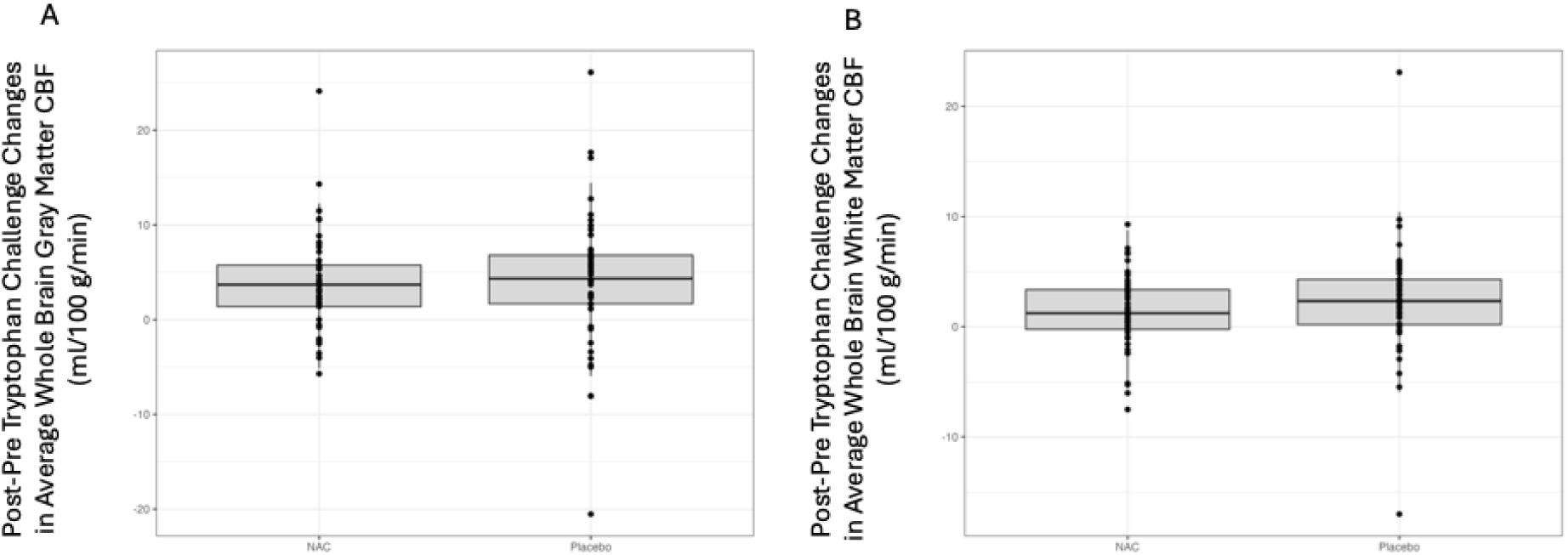
Effect of NAC on Tryptophan-Induced Changes in Gray and White Matter CBF. Footnote: ^a^The Panel A boxplot depicts the post-treatment minus pre-treatment difference in average whole brain gray matter CBF. There was a trend for a significant difference in difference between NAC and placebo pre-treatment on the baseline/post-treatment change in average whole brain gray matter CBF (avg-GM CBF; t=-2.15; p=0.078). The Panel B boxplot depicts the post-treatment minus pre-treatment difference in average whole brain white matter CBF. There was a significant difference in difference between NAC and placebo pre-treatment on the baseline/post-treatment change in average whole brain white matter CBF (avg-WM CBF; t=-2.151; p= 0.037). ALT text: The two boxplots depict the effect of NAC and placebo pre-treatment on changes in average whole brain gray and white matter cerebral bool flow.

<u>^1^H-MRS</u>. There were no significant treatment effects on glutamate (p=0.49) or glutathione (p=0.1).

<u>DTI</u>. There was a trend for a significant treatment effect for the average fractional anisotropy (FA) measure (t=-1.95; p=0.06), with participants treated with placebo showing increased post-treatment versus baseline change in FA.

**Table 2:** Total Gray and White Matter Arterial Spin Labeling (ASL) values.

| Total Gray Matter ASL (ml/100g/min) |  |  |  |  |
| --- | --- | --- | --- | --- |
| Timepoint | NAC |  | Placebo |  |
|  | mean | ± SD | mean | ± SD |
| Baseline | 49.9** | 11.0 | 52.5*** | 10.3 |
| 120 minutes | 54.4* | 12.0 | 57.3*** | 12.2 |
| Total White Matter ASL (ml/100g/min) |  |  |  |  |
| Timepoint | NAC |  | Placebo |  |
|  | mean | ± SD | mean | ± SD |
| Baseline | 18.3** | 4.4 | 18.8*** | 4.8 |
| 120 minutes | 19.7* | 4.3 | 21.2*** | 5.9 |

\*: n=51; \*\*: n=53; \*\*\*: n=54

### Cognitive Measures

There was a significant treatment effect, in which participants pre-treated with NAC did not exhibit the worsening of the MATRICS composite score with TRYP that was observed in the placebo-pre-treated group (see Supplemental Table 2 and Figure 3; t=2.07; p=0.04). There was also a significant effect of NAC treatment for the spatial working memory test: WMSIII-SS (t=2.41; p=0.02); performance on the WMSIII-SS improved in participants treated with NAC, whereas it worsened in those treated with placebo. There were no significant effects for any of the other individual MCCB subtests (all p values > 0.25).

### Clinical Measures

There were no significant treatment effects for any of the clinical outcome measures (see Supplementary Table 3; all p values ≥ 0.18).

### KMO SNP Measures

We examined the effect of two SNPs: rs2275163 and rs1053230 on those measures for which there had been a significant or trend effect of NAC pre-treatment: kynurenine and KYNA plasma levels; avg-WM and avg-GM CBF; and the MCCB Composite score and WMS-III Spatial Span measures. The number of participants for each rs2275163 genotype were: C/C: 28; C/T: 24; T/T: 3; and for each rs1053230 genotype: C/C; 44; C/T:10; and T/T: 1. There was a trend for NAC compared to placebo pre-treatment to reduce the TRYP-induced increase in plasma KYNA to a greater extent in those participants with the rs2275163 C/C genotype than those with either the C/T or T/T genotypes (see Supplementary Figure 3; t=1.71; p=0.095). The p values for all the other analyses were > 0.20). The were no significant or trend effects for the rs1053230 C risk allele (all p values > 0.20).

**Figure 3:**
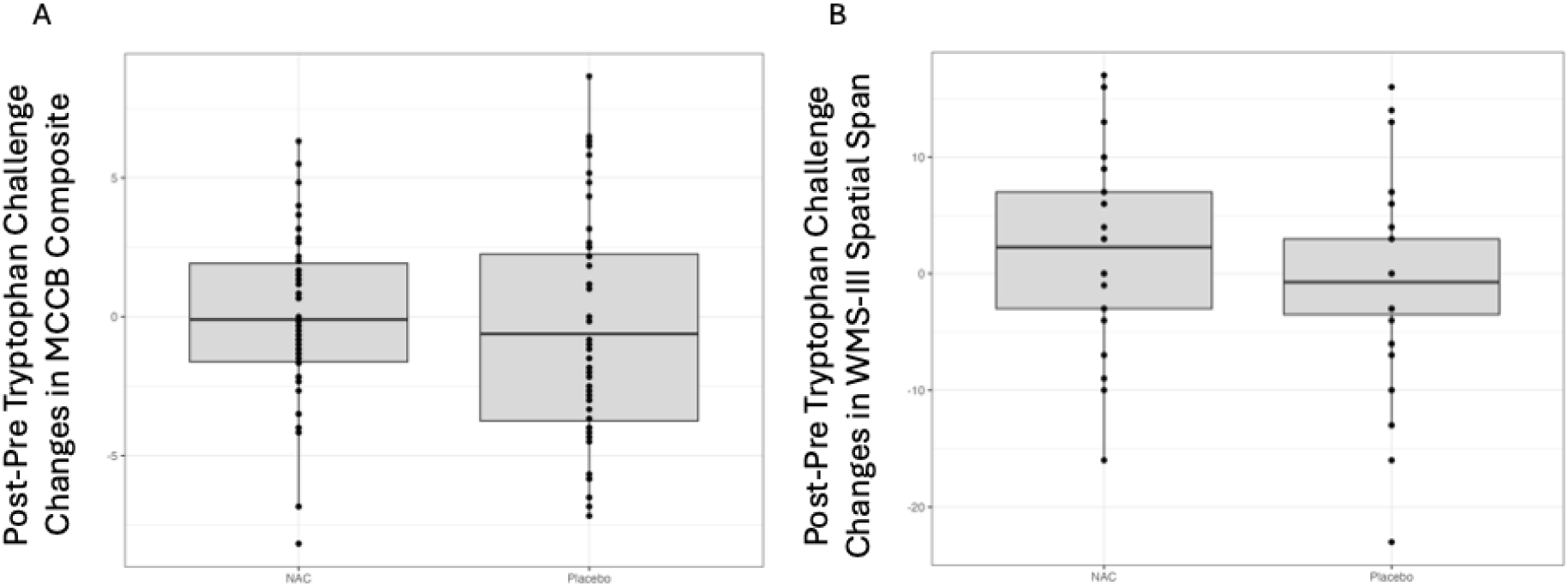
Effect of NAC on Tryptophan-Induced Changes in Cognition. Footnote: ^a^The Panel A boxplot depicts the post-treatment minus pre-treatment difference in the MCCB composite score. There was a significant difference in difference between NAC and placebo pre-treatment on the MATRICS composite score (t=2.07; p=0.04). The Panel B boxplot depicts the post-treatment minus pre-treatment difference in the Wechsler Memory Scale-III Spatial Span Working Memory test (WMSIII-SS). There was a significant difference in difference between NAC and placebo pre-treatment on the WMSIII-SS (t=2.41; p=0.02). ALT text: The two boxplots depict the effect of NAC and placebo pre-treatment on changes in the MCCB composite score and the Wechsler Memory Scale-III Spatial Span Working Memory test.

### Safety Assessments

There was one serious adverse effect (SAE): a participant was hospitalized overnight for suicidal ideation; the SAE was considered not study related, as it occurred prior to treatment randomization. There were twelve non-serious adverse events: 7 with placebo pre-treatment (nausea: 2, vomiting: 2; dizziness: 2; and anxiety: 1) and 5 with NAC pre-treatment (nausea: 2; vomiting: 2; and dizziness: 1).

## Discussion

The primary study aims were to examine whether high-dose NAC attenuates the TRYP-induced elevation of peripheral KYNA and the adverse effects associated with increased KYNA on selected measures of brain function. We found that pre-administration of NAC led to a significant reduction in plasma kynurenine and KYNA; diminished the TRYP-induced elevations in resting brain CBF in white matter (with a trend effect for gray matter); and mitigated the worsening of the MATRICS composite score and improved performance on the WMS-III Spatial Span.

The demonstration that high-dose NAC significantly decreases the conversion of TRYP to KYNA provides strong proof of principle evidence that KAT II inhibition can decrease production of KYNA in clinical populations. The observed effect was most pronounced 4 hours after TRYP administration, though initial effects were seen after 2 hours. The time course of the NAC effect on the conversion of kynurenine to KYNA is consistent with the results from our previous study, which demonstrated that peak serum levels of kynurenine and KYNA occur 90-150 minutes after TRYP administration and remain elevated for 4-6 hours [25].

NAC pretreatment reduced the TRYP-induced increase in CBF in PSz. The significant effect of high-dose NAC pretreatment on avg-WM CBF (with a trend-level effect on avg-GM CBF) suggests that NAC was effective in blocking the conversion of kynurenine to KYNA within the CNS. Although there was only a trend-level effect of NAC on average-GM CBF, there were 5 individual cortical regions, for which there was a medium or greater Cohen’s d effect size. Moreover, the majority of cortical regions showed that NAC reduced the increase in CBF following TRYP administration compared to the placebo arm. In light of the time course of NAC effects on KYNA levels, we might have observed a more pronounced effect on ASL if the scans had been obtained when NAC was observed to maximally exert its effect on the conversion of kynurenine to KYNA, i.e., 3-4 hours after TRYP administration. Finally, the lack of effect of high-dose NAC pretreatment on brain GSH or glutamate levels suggests that the effect of NAC on CBF was not due to its antioxidant properties or the modulation of glutamate release.

The ultimate goal of interfering with the formation of KYNA is to lower the amount of brain KYNA available to antagonize α-7 nicotinic and NMDA glutamate receptors and thereby reduce the adverse impact of these interactions on cognitive function. In the current study, we found that pre-treatment with high-dose NAC prevented the worsening of cognitive function following TRYP administration. Furthermore, participants who received NAC also demonstrated a significant improvement in their performance on a spatial working memory test. These results provide compelling evidence that KAT II inhibition may be a viable approach for the treatment of cognitive impairments in PSz.

In the context of exploratory analyses examining whether the rs2275163 and rs1053230 SNPs impacted the effects of NAC on any of the outcome measures, we found a trend effect of the rs2275163 C risk allele for KYNA. Although not statistically significant, there was a moderate effect size (Cohen’s d=0.51) for the analysis, which showed that participants with the CC genotype had a greater response to NAC pre-treatment than those with either the CT or TT genotypes. A possible explanation of the observed effect is that the CC genotype is associated with decreased KMO activity in PSz [17], which would have the effect of shifting the metabolism of kynurenine from 3-OH kynurenine to KYNA. However, high-dose NAC inhibits KAT II, which would counteract the effect of decreased KMO expression. If the mediating effect of the CC genotype on NAC reduction of plasma KYNA is confirmed in larger studies, then this would provide further support for the utility of a KAT II inhibitor in the treatment of PSz.

NAC pretreatment had no significant effects on any of the symptom measures, which suggests that NAC neither improved nor worsened symptoms over the course of the challenge day. In addition, NAC pretreatment, compared to placebo, was not associated with an increased incidence of adverse events.

In summary, the study results provide strong evidence for the potential utility of a KAT II inhibitor to attenuate the conversion of kynurenine to KYNA and reverse the adverse effects of elevated KYNA on CBF and cognitive function. The development of specific KAT II inhibitors is required to assess the clinical relevance of this pharmacological approach for the treatment of people with schizophrenia and beyond.

## Data Availability

All data produced in the present study are available upon reasonable request to the authors.

## Author Contributions

SMH took lead in overseeing data analysis and manuscript drafting; DLK assisted with obtaining funding for the project and assisted with collection, storage and analysis of the blood draws, including the genetics analysis; YP took lead in data analysis and generation of figures; SC was the primary statistician for the project and oversaw all data analyses; FB was the pharmacist overseeing preparation and administration of the TRYP and NAC slurries; DAG was the study physician providing oversight and assistance with patient vitals and adverse events; JMG is a cognitive neuroscientist providing expertise in the deployment and analysis of measures of cognitive function; KVS provided essential expertise in the methods to measure and analyze the KP metabolites, and the genetics analysis (and assisted with drafting Methods sections; BMA provided essential expertise and time to perform the processing and analysis of the ASL cerebral blood flow measures; PK provided oversight and expertise in the imaging analyses, especially assisting with the DTI analyses (he was essential in drafting sections of the Methods); SAW provided oversight and expertise in the imaging analyses, especially assisting with the MRS analyses; LMR provided oversight and expertise in the imaging analyses, especially assisting with the MRS analyses (she also was essential in obtaining funding for the project and drafting sections of the Methods); RS was the PI for the 2P50MH103222 grant that funded this research, playing key roles in overseeing data analysis and drafting sections of the manuscript; RWB was senior author and played key roles in obtaining funding for the project, overseeing the design and conduct of the study, and manuscript drafting.

