## Supplemental Material for "N-Acetylcysteine Reduces Tryptophan-induced Abnormalities in People with Schizophrenia"

**Supplementary Methods**

**Kynurenine and KYNA Measurements:** Peripheral venous blood was collected 150 minutes before (baseline fasting levels) and 60, 140 and 240 minutes after the ingestion of TRYP. The blood samples were collected in EDTA-containing tubes and centrifuged immediately (within 30 minutes after the blood draw). After centrifugation, the supernatant plasma was removed and stored at -80^o^C until analysis. Following thawing and dilution of the plasma sample (1:2 v/v in ultrapure water) on the day of the assay, 200 µl of the sample were deproteinized by the addition of 50 µl of 6% perchloric acid. Samples were thoroughly mixed, centrifuged (16,000 x g, 15 minutes), and 20 µl of the resulting supernatant were then applied to a 3-µm ReproSil C18 column (100 mm x 4 mm; Dr. Maisch GmbH, Ammerbuch, Germany). Kynurenine and KYNA were isocratically eluted using a mobile phase containing 50 mM sodium acetate and 5% acetonitrile (pH adjusted to 6.2 with glacial acetic acid) at a flow rate of 0.5 ml/min. Using post-column derivatization with 500 mM zinc acetate, delivered at a flow rate of 0.1 ml/min, the two KP metabolites were measured in the eluate using fluorimetric detection [kynurenine: excitation 365 nm, emission 480 nm; KYNA: excitation 344 nm, emission 398 nm; Perkin-Elmer series 200; Waltham, MA, USA]. The retention times of kynurenine and KYNA were ~7 minutes and ~13 minutes, respectively.

**SNP Genotyping:** Genomic DNA was extracted from human whole blood collected in PAXgene Blood DNA Tubes using the PAXgene Blood DNA Kit (QIAGEN/PreAnalytiX, Cat. No. 761133) according to the manufacturer’s instructions. A total of 10 ng of purified genomic DNA was used for genotyping. We focused primarily on the *KMO* intronic variant rs2275163 (C>T) and rs1053230 (C>T). Both SNPs have minor allele frequencies ≥ 0.15 in European American and African American/mixed populations. The corresponding TaqMan Genotyping Assays (Applied Biosystems) were: rs2275163 (Assay ID C_16183814_10) and rs1053230 (Assay ID C_8856260_10). Genotyping was performed using TaqMan SNP Genotyping Assays employing the 5’ nuclease real-time PCR method. Each assay includes sequence-specific forward and reverse primers to amplify the target region containing the polymorphism. Allelic discrimination was carried out using two TaqMan minor groove binder (MGB) probes: a probe labeled with VIC® dye to detect the reference allele (Allele 1) and a probe labeled with FAM™ dye to detect the alternate allele (Allele 2). Fluorescent signals were measured during PCR amplification, and alleles were automatically assigned using the instrument’s allelic discrimination software. Standard internal controls and quality-control metrics were applied to ensure assay performance. Based on previous findings (1), we compared phenotypes across 2 genotype groups (homozygous CC vs. combined CT/TT genotypes).

**MRI Assessments:**

Structural. The following parameters were used to collect the structural scans: T1-weighted MPRAGE, TR/TE = 2400/2.2 ms, flip angle =8°, spatial resolution = 0.8 mm x 0.8 mm x 0.8 mm with 208 slices per slab. All data were preprocessed using the HCP minimal processing pipeline, which included FreeSurfer analysis including quantification of regional cortical thickness and subcortical gray matter volumes (11).

Arterial Spin Labeling (ASL). The following parameters were used to collect pseudo-continuous ASL data: TR/TE = 4000/37 ms, 2.3 mm × 2.3 mm × 2.6 mm with 58 axial slices, post-label delay = 1.7 s, labeling duration=1.65 s. For normalization purposes, a volume of M_0_ image was acquired without background suppression, and smoothed with a 5 mm Gaussian-kernel, consistent with recent recommended processing guidelines (2). Spatial regularization, motion correction and partial volume corrections using a spatially regularized method (3) were performed using FSL software. Partial volume estimates for gray matter, white matter and cerebrospinal fluid were obtained from each participant’s T_1_-weighted structural image and transformed to the ASL native image space using a transformation matrix from the structural space. Partial volume corrected CBF maps were used to extract the whole-brain average CBF across all gray matter and across all white matter. The average CBF signals for 246 ROIs were extracted using the Brainnetome atlas.

Magnetic Resonance Spectroscopy (^1^H-MRS). The following parameters were used to collect ^1^H-MRS data: very short TE phase rotation STEAM with TR/TM/TE= 2000/10/6.5 ms, NEX=128 (metabolite) and 16 (water reference), 2500-Hz spectral width, 2048 complex points, and Δφ1=135°, Δφ2=22.5°, Δφ3=112.5°, and ΔφADC=0°) optimized for the detection of glutamate (4) and shown to have excellent reproducibility for glutamate and glutathione (5-7). Spectra were analyzed with the fully automated, standard curve-fitting software, LCModel (8) using a basis set that included alanine (Ala), aspartate (Asp), creatine (Cr), γ-aminobuytric acid (GABA), glucose (Glc), Glu, glutamine (Gln), glutathione (GSH), glycine (Gly), glycerophosphocholine (GPC), lactate (Lac), myo-Inositol (mI), N-acetylaspartate (NAA), N-acetylaspartylglutamate (NAAG), phosphocholine (PCh), phosphocreatine (PCr), phosphoroylethanolamine (PE), scyllo-Inositol (sI), and taurine (Tau), simulated in Gamma Visual Analysis (9). Spectra with SNR>10 and water FWHM <0.01ppm were included. Metabolites were referenced to water and presented in institutional units (IU) and only metabolites of good fits (%SD CRLBs< 20) were included. Spectroscopic voxels were tissue segmented using SPM12 and metabolites were corrected for CSF, gray, and white matter water content and metabolite and water relaxation times according to Gasparovic et al (10).

Diffusion Tensor Imaging (DTI). Diffusion-weighted imaging (DWI) data were collected using an expansion of the HCP protocol that consisted of 6 shells of *b*-values (*b*=600, 900, 1200, 1500, 1800, and 3000 s/mm^2) with 98 isotropically distributed diffusion-weighted directions per shell collected twice with a reversal of the phase encoding and readout gradients (anterior-to-posterior AP and posterior-to-anterior PA) to correct for spatial distortions. We also included twenty b=0 images interleaved within the acquisition. The data was collected using a multiband, echo-planar, spin-echo, T2-weighted sequence (TE/TR/Multiband Factor=97/4000 ms/4 with the FOV=200 mm) with an isotropic spatial resolution of 1.6 mm. Diffusion MRI data were preprocessed using the HCP Diffusion pipeline (11,12) that was combined with DESIGNER diffusion preprocessing tools, including advanced denoising, Gibbs ringing correction, and correction of EPI distortions (13). FA maps were obtained by fitting the diffusion tensor model using the FSL-FDT toolkit (14).

**Supplementary Table 1: Demographic Characteristics**

| **Variable** |  | |
| --- | --- | --- |
| Age (mean ± SD) | 40.2 | 9.3 |
| Sex |  | |
| Female | 16 (27.6%) | 27.6% |
| Male | 42 (72.4%) | 72.4% |
| Race |  | |
| White | 27 | 46.6% |
| Black | 27 | 46.6% |
| Other | 4 | 6.8% |

**Supplemental Table 2: Cognitive Measures**

|  | **NAC** | | **Placebo** | |
| --- | --- | --- | --- | --- |
| Timepoint | mean | ± SD | mean | ± SD |
| **MCCB Composite Score** | | | | |
| Baseline | 41.5^++^ | 8.3 | 42.2^+^ | 8.3 |
| 150 minutes | 42.2*** | 8.3 | 41.6* | 7.7 |
| **WMS-III Spatial Span** | | | | |
| Baseline | 40.2^+++^ | 10.7 | 43.4^++^ | 10.2 |
| 150 minutes | 43.4**** | 11.2 | 42.4**** | 10.8 |
| **Letter-Number Sequencing** | | | | |
| Baseline | 42.8^+++^ | 10.8 | 41.8^++^ | 10.9 |
| 150 minutes | 44.5**** | 12.1 | 43.0**** | 10.2 |
| **BACS Symbol Coding** | | | | |
| Baseline | 40.6^++^ | 14.3 | 40.6^++^ | 13.1 |
| 150 minutes | 42.0**** | 15.7 | 43.9*** | 14.3 |
| **Brief Visuospatial Memory Test-Revised** | | | | |
| Baseline | 44.3^++^ | 11.9 | 43.5^+^ | 11.6 |
| 150 minutes | 40.2**** | 11.3 | 40.2** | 9.8 |
| **Hopkins Verbal Learning Test-Revised** | | | | |
| Baseline | 39.2^+++^ | 9.2 | 40.1^++^ | 10.6 |
| 150 minutes | 38.4**** | 9.7 | 37.8**** | 10.8 |
| **CPT-Identical Pairs** | | | | |
| Baseline | 42.0^+++^ | 12.4 | 43.5^++^ | 11.8 |
| 150 minutes | 42.3*** | 10.9 | 42.7**** | 10.8 |

________________________

^*^: n=48; ^**^: n=49; ^***^: n=50; ^****^: n=51; ^+^: n=53; ^++^: n=54; ^+++^: n=55

MCCB, MATRICS Consensus Cognitive Battery; WMS, Weschler Memory Scale; BACS, Brief Assessment of Cognition in Schizophrenia; CPT, Continuous Performance Test

**Supplemental Table 3: Clinical Measures**

| **BPRS Total Score** | | | | |
| --- | --- | --- | --- | --- |
|  | **NAC** | | **Placebo** | |
| Timepoint | mean | ± SD | mean | ± SD |
| Baseline | 34.5^+^ | 7.2 | 34.4^++^ | 6.3 |
| 190 minutes | 33.8* | 7.3 | 34.2*** | 6.4 |
| **BPRS Positive Symptom Score** | | | | |
| Baseline | 7.5^+^ | 3.2 | 7.8^++^ | 3.4 |
| 190 minutes | 7.5* | 3.3 | 7.8*** | 3.5 |
| **SANS Total Score** | | | | |
| Baseline | 23.1^+^ | 8.0 | 23.1^++^ | 9.6 |
| 190 minutes | 22.6* | 8.5 | 23.1*** | 9.6 |
| **CDS Total Score** | | | | |
| Baseline | 1.8^++^ | 2.5 | 1.3^++^ | 1.8 |
| 190 minutes | 1.6* | 2.5 | 1.0*** | 1.6 |
| **CGI Global Severity** | | | | |
| Baseline | 4.0^+^ | 0.8 | 3.9^++^ | 0.8 |
| 190 minutes | 4.0** | 0.8 | 3.9*** | 0.8 |

________________________

^*^: n=52; ^**^: n=53; ^***^: n=54; ^+^: n=56; ^++^: n=57

BPRS, Brief Psychiatric Rating Scale; SANS, Scale for the Assessment of Negative Symptoms; CDS, Calgary Depression Scale; CGI, Clinical Global Inventory

**Supplementary Figure 1 – Consort Flow Diagram**

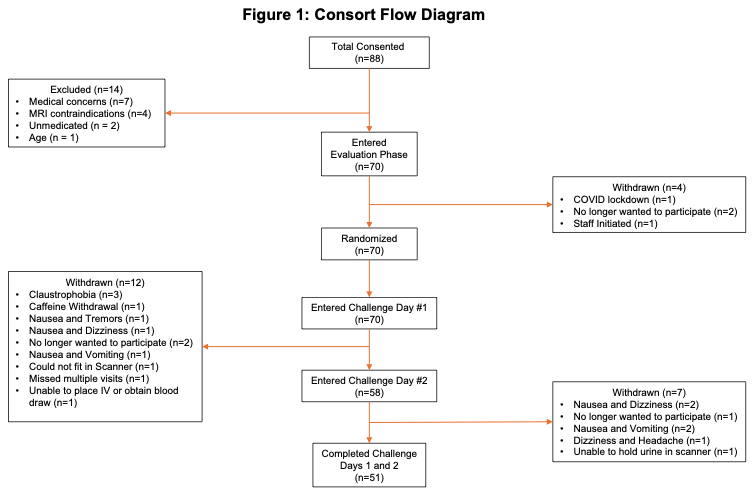

**Supplementary Figure 2 - Standardized Effect Sizes for the Treatment-Related Differences-in-Differences across the Brainnetome Atlas Cortical ROIs^a^**

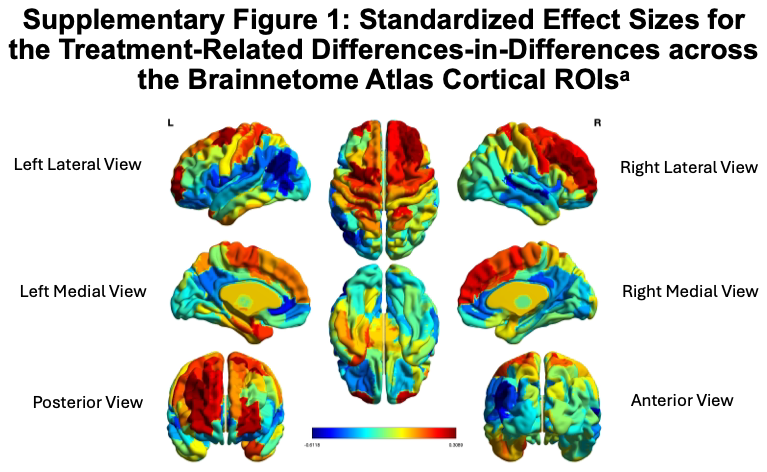

**Supplementary Figure 3. The Change in KYNA Plasma Levels by KMO SNP rs2275163 Genotype^a^**

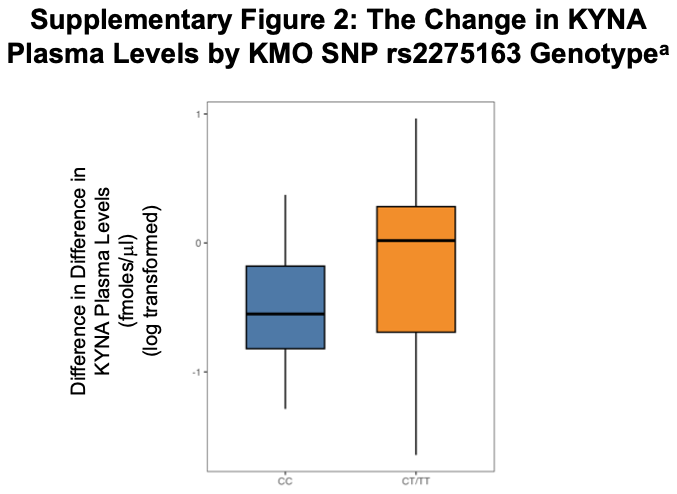

**Figure Legends:**

**Supplementary Figure 1:**

Title: CONSORT Flow Diagram

Footnote: N/A

ALT text: CONSORT diagram showing participant flow through the study.

**Supplementary Figure 2:**

Title: Standardized Effect Sizes for the Treatment-Related Differences-in-Differences across the Brainnetome Atlas Cortical ROIs

Footnote: Cortical map of regional standard effect sizes for difference in difference changes in cerebral blood flow following NAC or placebo pre-treatment. The regions in which the change in cerebral blood flow is greater with NAC pre-treatment are reflected in the scale orange and red colors and the regions in which the change in cerebral blood flow is greater with placebo pre-treatment are reflected in the scale colors from blue to yellow.

ALT text: A cortical map of regional standard effect sizes for difference in difference changes in cerebral blood flow following NAC or placebo pre-treatment. The regions in which the change in cerebral blood flow is greater with NAC pre-treatment are reflected in the scale orange and red colors and the regions in which the change in cerebral blood flow is greater with placebo pre-treatment are reflected in the scale colors from blue to yellow.

**Supplementary Figure 3:**

Title: The Change in KYNA Plasma Levels by KMO SNP rs2275163 Genotype

Footnote: The boxplots represent the difference in difference following NAC or placebo pre-treatment for plasma KYNA levels grouped according to KMO SNP rs2275163 genotype: CC or CT/TT. There was a trend for NAC to reduce the TRYP-induced increase in plasma KYNA compared to placebo to a greater extent in those participants with the rs2275163 C/C genotype than those with either the C/T or T/T genotypes (t=1.71; p=0.095).

ALT text: The boxplot depicts the effect of NAC and placebo pre-treatment on changes in plasma KYNA levels based on participant KMO SNP rs2275163 genotype.
